# Acquisition and carriage dynamics of fluoroquinolone resistant Enterobacteriaceae at individual and household levels

**DOI:** 10.1101/2020.10.29.20222364

**Authors:** Patrick Musicha, Andrew J. Stewardson, Yin Mo, Jascha Vervoort, Niels Adriaenssens, Samuel Coenen, Maciek Godycki-Cwirko, Anna Kowalczyk, Christine Lammens, Surbhi Malhotra-Kumar, Herman Goossens, Stephan Harbarth, Ben S. Cooper

## Abstract

Carriage dynamics of drug-resistant bacteria, especially within households, are poorly understood. This limits the ability to develop effective interventions for controlling the spread of antimicrobial resistance in the community. Two groups consisting of: (i) patients with urinary tract infection requiring antimicrobial treatment; and (ii) patients who were not prescribed antimicrobial treatment were prospectively recruited at three European sites: Antwerp (Belgium), Geneva (Switzerland) and Lodz (Poland). Each index patient and up to three additional household members provided faecal samples at baseline, completion of antimicrobial therapy (or 7-10 days after the first sample for the non-exposed) and 28 days after the second sample. We analysed household-level and individual-level fluoroquinolone resistant Enterobacteriaceae (FQR-E) acquisition and carriage data using Bayesian multi-state Markov models. At the individual level, we estimated a median baseline FQR-E acquisition rate of 0.006 (95%*CrI* = [0.004, 0.01]) per day, and a median duration of carriage of 24.4 days (95% CrI=[15.23,41.38]). Nitrofurantoin exposure was associated with a reduced rate of FQR-E acquisition (HR=0.28, 95%CrI=[0.14,0.56]), while fluoroquinolone exposure had no clear association with rates of FQR-E acquisition (HR=1.43, 95% CrI=[0.81,2.53]) at individual level. There was evidence that rates of FQR-E acquisition varied by site, and coming from Lodz was associated with a higher acquisition rate (HR=3.56, 95% CrI=[1.92, 6.34]). Prolonged duration of carriage was associated with exposure to fluoroquinolone or nitrofurantoin during the study, use of any antimicrobial agent in the prior 12 months and travel to endemic regions. At household level, we found strong evidence of positive association between FQR-E acquisition and fluoroquinolone exposure (HR=3.43, 95% CrI=[1.51,7.74]). There was weak evidence of negative association between FQR-E acquisition and nitrofurantoin exposure (HR=0.42, 95%CrI=[0.12, 1.24]. Similar to the individual level, carriage duration was also associated with antimicrobial exposure at the household level. Our study has identified within household contacts as an important route for FQR-E transmission and highlights the need for prioritising household focused interventions to control FQR-E spread.

## Introduction

Fluoroquinolones are among the the most commonly used antimicrobial agents worldwide and are effective against several common bacterial infections including intra-abdominal, urinary tract, pneumonia and soft tissue infections [1, 2, 3, 4]. The clinical application of fluoroquinolones is however, being threatened by the global spread of multi-drug resistant (MDR) bacteria [5, 6]. Not only have fluoroquinolones been classified as one of the “Watch antibiotics” by the World Health Organization for their high potential to influence the development of antimicrobial resistance (AMR)[7], there is growing evidence from various settings that increased use of fluoroquinolones has been accompanied by rapid emergence and spread of fluoroquinolone-resistant Enterobacteriaceae (FQR-E) [8, 2, 6, 9]. Furthermore, FQR-E are often co-resistant to other broad-spectrum antimicrobials such as third generation cephalosporins (3^*rd*^GC), leading to increased dependence on carbapenems [8, 2, 6, 9]. Preserving the effectiveness of fluoroquinolones would therefore help to ensure availability of alternative treatment options for infections caused by bacteria with either intrinsic or fast-spreading acquired resistance to other commonly used broad-spectrum antimicrobials. [10]

Antimicrobial use is a major driver of emergence and spread of AMR [11, 8]. While effects of antimicrobial exposure on carriage dynamics of drug-resistant Enterobacteriaceae have been studied at the hospital level with detailed individual-level data [12], comparable analyses in community settings have been lacking. Both ecological and individual level factors contribute toward the spread of MDR bacteria in the community [11, 13, 14, 15, 10]. A good understanding of these complex dynamics is important for the development of effective interventions to combat the spread of AMR and for quantifying the likely effect of changing patterns of antimicrobial use. We would like to know how quickly drug-resistant bacteria in various settings are acquired and cleared, how important different routes of transmission are, and what effect antimicrobial use and other factors have on these processes. Mathematical models, when fitted to high quality individual-level observational data, are potentially valuable tools for quantifying these dynamics and for predicting the impact of interventions [16]. Previous studies investigating acquisition and carriage dynamics of drug-resistant Enterobacteriaceae have mainly focused on ESBL-E and carbapenemase producing Enterobacteriaceae (CPE)[13, 17, 12, 18]. Furthermore, only a few of the ESBL-E studies have described transmission of drug-resistant bacteria within households, and until now analysis informed by dynamical models accounting for antimicrobial exposure has been lacking [19]. Our understanding of the role of household contacts in the transmission of drug-resistant bacteria, and in particular of FQR-E, therefore remains limited. Unlike ESBL and carbapenem resistance, the main genetic mechanism of high-level FQR is chromosomal mutation [20, 21, 4]. While FQR mutants may emerge within hosts during fluoroquinolone treatment, the spread of FQR-E is largely driven by the expansion of a few successful MDR clones such as the *E. coli* ST131 H30Rx [22, 23]. Spread of such clones is mostly through person-to-person transmission, which could be enhanced within households due to the shared environment and proximity of household members [19]. Here, we investigated transmission dynamics of FQR-E at individual and household levels by fitting multi-state Markov models on longitudinal data from a multi-site European cohort. We quantified the effects of antimicrobial (fluoroquinolone and nitrofurantoin) exposure on the acquisition and carriage of FQR-E while accounting for other covariates.

## Results

### Characteristics of study participants

Five hundred and ninety-one individuals from three European sites (Antwerp (Belgium), Geneva (Switzerland) and Lodz (Poland)) participated in a cohort study between January 2011 and August 2013. Faecal samples were provided at at least two sampling time points by 590 (99.8%) participants and at up to three sampling time points by 524 (88.7%). One participant was sampled only once and was excluded from further analysis. In this cohort, 144 (24.4%) participants were index patients who had urinary tract infection (UTI) and required antimicrobial therapy. Sixty-nine (47.9%) of these index patients were treated with fluoroquinolone and 75 (52.1%) were treated with nitrofurantoin. Four hundred and forty six (75.6%) participants were exposed to neither of these two antimicrobial agents (Table 1). Eighty-eight (14.9%) participants were already colonised at the first sampling time point, 98 (16.6%) were identified as colonised at the second sampling time point and 83 (15.8%) at the third sampling time point (Figure 1).

**Table 1:** Summary of participants’ and household characteristics and covariates at three sampling time points (TP1, TP2 and TP3). * Use of any antimicrobial within prior 12 nonths

| Char | Antwerp |  |  | Geneva |  |  | Lodz |  |  |
| --- | --- | --- | --- | --- | --- | --- | --- | --- | --- |
|  | TP1 | TP2 | TP3 | TP1 | TP2 | TP3 | TP1 | TP2 | TP3 |
| Samples, n(%) | 144 (100.0) | 144 (100.0) | 123 (85.4) | 233 (100.0) | 233 (100.0) | 210 (90.1) | 213 (100.0) | 213 (100.0) | 191 (89.7) |
| Mean age | 28.6 |  |  | 30.6 |  |  | 41.5 |  |  |
| Sex (F) | 80 (55.6) | 80 (55.6) | 66 (53.4) | 131 (56.2) | 131 (56.2) | 120 (57.1) | 121 (56.8) | 121 (56.8) | 110 (57.6) |
| *Households* |  |  |  |  |  |  |  |  |  |
| Two-member | 29 (56.9) | 29 (56.9) | 23(53.8) | 49 (55.1) | 49 (55.1) | 44 (57.9) | 90 (92.8) | 90 (93.8) | 90 (93.8) |
| Three-member | 14 (27.5) | 14 (33.3) | 14 (27.5) | 27 (30.3) | 27 (30.3) | 22 (28.9) | 3 (3.1) | 3 (3.1) | 3 (3.1) |
| Four-member | 8 (15.7) | 8 (15.7) | 5 (11.9) | 13 (14.6) | 13 (14.6) | 10 (13.2) | 4 (4.1) | 3 (3.1) | 3 (3.1) |
| *Antimicrobial exposure* |  |  |  |  |  |  |  |  |  |
| No exposure | 125 (86.8) | 125 (86.8) | 107 (87.0) | 180 (77.3) | 180 (77.3) | 159 (75.7) | 141 (66.2) | 141 (66.2) | 130 (68.1) |
| Nitrofurantoin | 16 (11.1) | 16 (11.1) | 13 (10.6) | 21 (9.0) | 21 (9.0) | 21 (10.0) | 38 (17.8) | 38 (17.8) | 34 (17.8) |
| Ciprofloxacin | 3 (2.1) | 3 (2.1) | 3 (2.4) | 32 (13.7) | 32 (13.7) | 30 (14.3) | 34 (16.0) | 34 (16.0) | 27 (14.1) |
| Antimicrobial use* | 24 (17.8) | 24 (17.8) | 22 (19.0) | 76 (32.9) | 76 (32.9) | 68 (32.7) | 75 (35.7) | 75 (35.7) | 67 (35.6) |
| Travel | 22 (15.3) | 22 (15.3) | 22 (17.9) | 33 (14.2) | 33 (14.2) | 31 (14.8 ( | 2 (0.9) | 2 (0.9) | 1 (0.5) |

**Figure 1:**
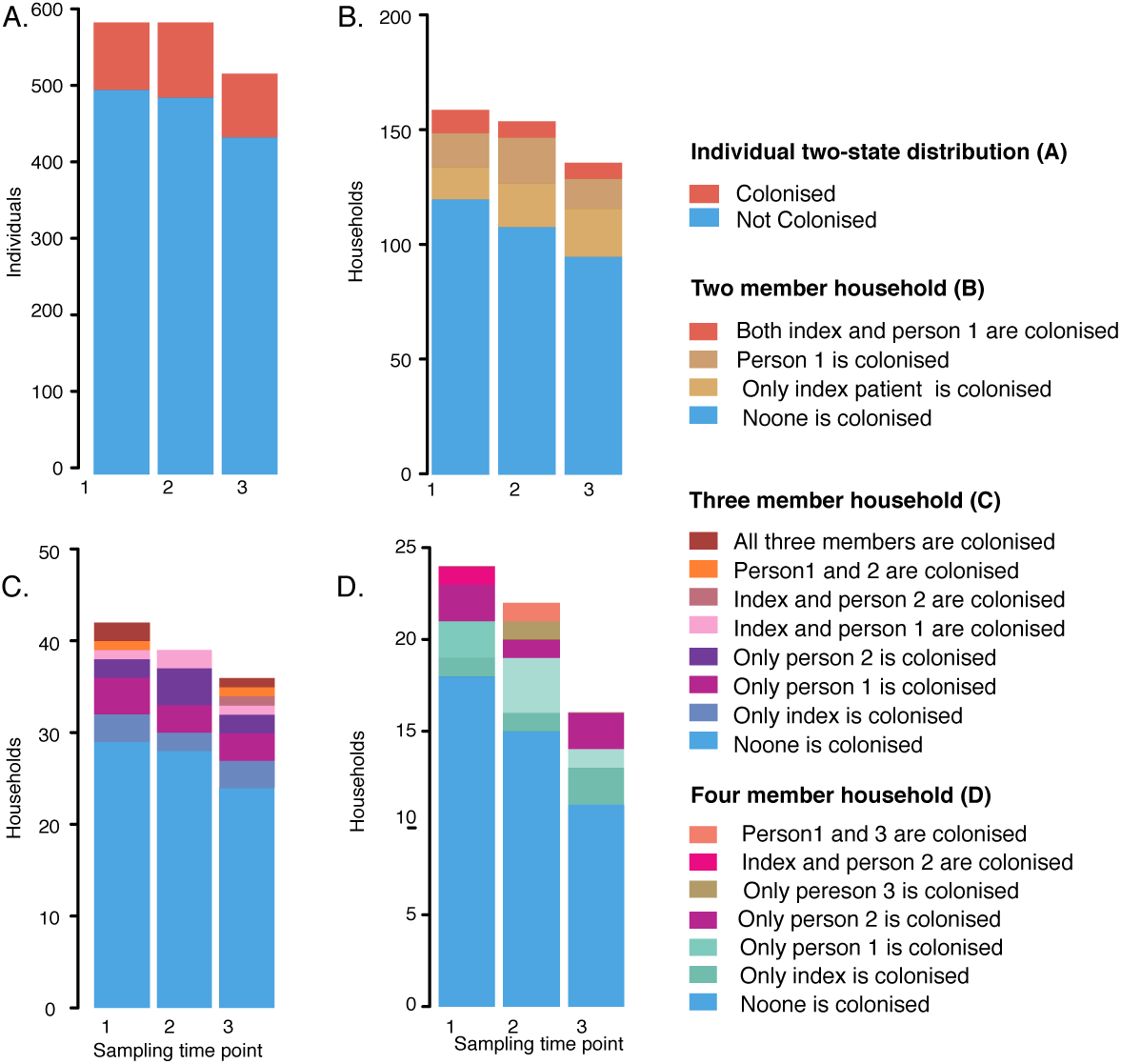
Distributions of individuals and households by colonisation status at three sampling time-points. Figure 2 (A) shows the number of individuals by their colonisation status at sampling time-points one, two and three. Figures 1(C-D) shows the number of two-member, three-member and four-member households by colonisation status, defined by the number of household members colonised, at sampling time-points one, two and three.

A total of 76 new acquisition events were detected between sampling time points one and three (46 at sampling time point two and 30 at sampling time point three). There were 85 decolonisation events during follow up which were detected at sampling time points two (36 events) and three (49 events). Of the 590 subjects with at least two samples, 144 (24.4%) came from Antwerp, 233 (39.5%) from Geneva and 213 (36.1%) from Lodz (Table 1). The 590 participants came from 259 households including 168 (64.9%) two-member households, 44 (17.0%) three-member households, 25 (9.7%) four-member households, while 22(8.5%) participants came from single-member households (Table 1).

### Acquisition and carriage dynamics of FQR-E at the individual level

We initially fitted a two-state individual level Markov model to the data for all the 590 subjects who had provided at least two faecal samples. Estimates from this model gave a mean baseline rate for acquiring FQR-E among individuals of 0.0061 (95% credible interval (CrI)=[0.004, 0.01]) cases per day (the reciprocal of this estimate gives a mean time to acquisition of 164 days). Individuals were cleared of colonisation at a baseline rate of 0.041 (95% CrI=[0.025, 0.067]) cases per day (equivalent to a mean time to clearance of 24.4 days in the absence of antimicrobial exposure). The estimated rate of FQR-E colonisation was higher amongst individuals who had been exposed to fluoroquinolone during the study, although uncertainty was large (Hazard ratio (HR)=1.43, 95% CrI=[0.81, 2.53]). Residing in Lodz was also associated with higher rates of colonisation (HR=3.45, 95% CrI=[1.92,6.34]). Conversely, exposure to nitrofurantoin during the study was associated with lower rates of FQR-E acquisition (HR=0.29, 95%CrI=[0.14,0.56]; Figure 2A(i)). FQR-E clearance within individuals was negatively associated with exposure to fluoroquinolone (HR=0.34, 95%CrI=[0.17,0.66]) or nitrofurantoin (HR=0.27, 95% CrI=[0.11, 0.61]) indicating prolonged carriage in individuals who had been exposed to these antimicrobial agents. Use of any antimicrobial agent in the prior 12 months or travel to endemic regions were also associated with reduced clearance rates (Figure 2A(ii)). FQR-E decolonisation was estimated to be faster in individuals from Lodz compared to those from Antwerp (HR=2.56, 95%CrI=[1.40, 4.60]), with a baseline duration of carriage estimated to be just about 10 days.

**Figure 2:**
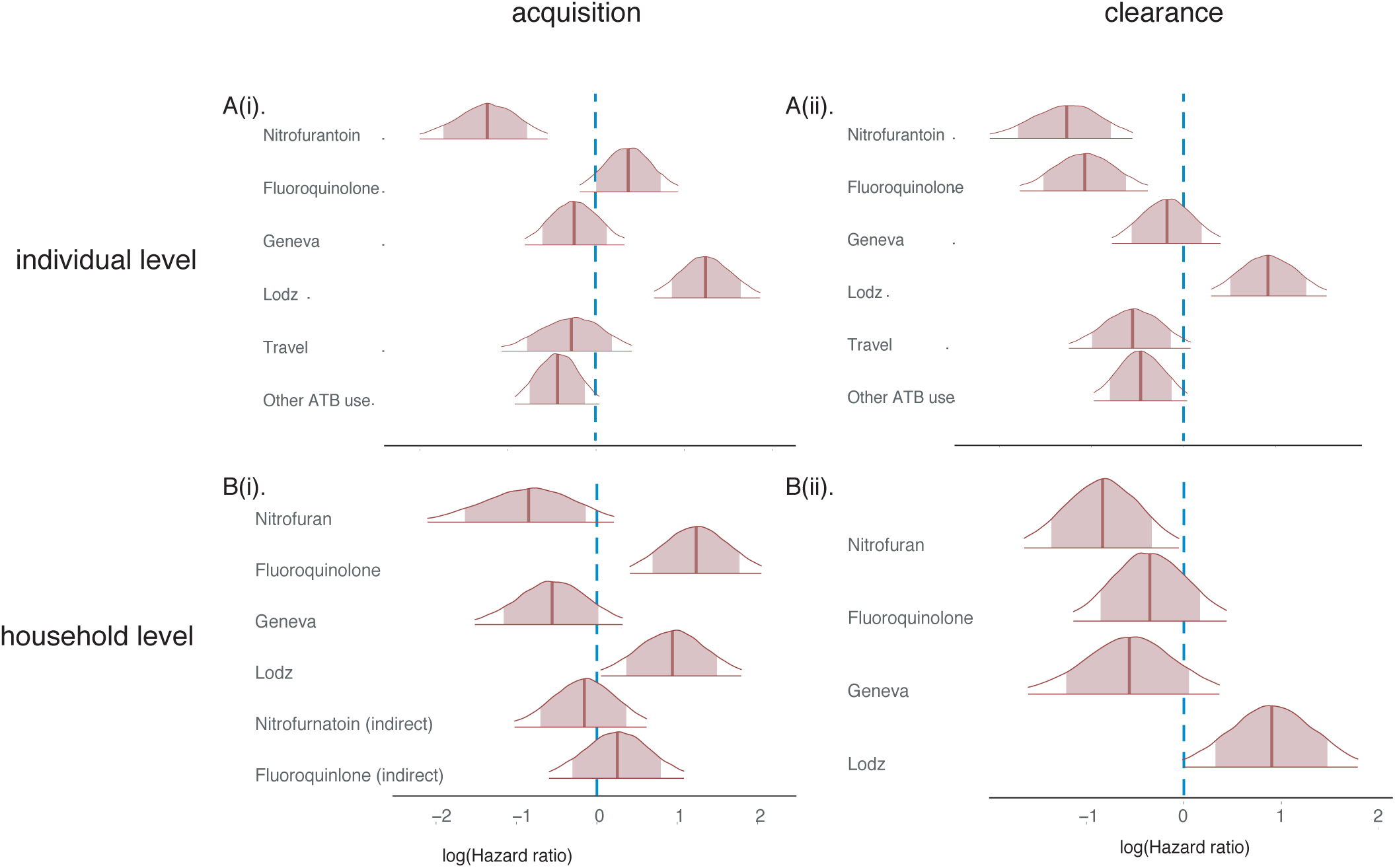
Effects of covariates on acquisition and carriage of FQR-E at individual level and within households. Panel A (i) and A(ii) show the posterior distributions of effects (hazard ratios on a log scale) of exposure to antimicrobial agents (nitrofurantoin or fluoroquinolone), location (Geneva and Lodz, with Antwerp as the baseline location), travel to AMR endemic regions and use of any antimicrobial agents in prior one year, on the rate of acquisition and decolonisation of FQR-E, respectively, at the individual level. Panels B(i) and B(ii) show the posterior distribution of effects antimicrobial exposure (nitrofurantoin and fluoroquinolone) and effect of location on FQR-E acquisition and decolonisation within households. Panel B(i) shows the effects of the covariates on rate of acquiring FQR-E within a household whereas B(ii) shows the effects of antimicrobial exposure and location on loss of carriage within household.

### Household level FQR-E acquisition dynamics

The mean baseline rate of FQR-E acquisition at the household level, i.e. the rate of a household transitioning from a state of having no colonised member to a state of having one colonised member was *λ*_*h,C*_ = 0.008, (95%CrI=[0.003, 0.02]). The baseline rate of a household member acquiring FQR-E from within the household was *λ*_*h,H*_ = 0.012, 95%CrI=[0.002, 0.04]. This means that in a household where at least one member was already colonised, it would take an average of 83 days for another individual to acquire FQR-E as compared to 125 days for an individual in a household where no member was known to be previously colonised. We investigated the effect of antimicrobial exposure on household level acquisition and clearance of FQR-E adjusted for household location. The household level analysis found strong evidence of an increased rate of FQR-E acquisition in fluoroquinolone exposed individuals (HR=3.43, 95%CrI=[1.51,7.74]) (Figure 2B(i)). In contrast, rates of FQR-E acquisition were not substantially elevated among individuals not exposed to fluoroquinolone within fluoroquinolone exposed households (HR=1.28, 95%CrI=[0.55, 2.95]; Figure 2B(i)). There was weak evidence of a negative association between nitrofurantoin exposure and FQR-E acquisition in exposed individuals (HR=0.42, 95%CrI=[0.12, 1.24]; Figure 2B(i)). As with fluoroquinolones, there was little evidence of indirect effects of nitrofurantoin exposure; i.e. the FQR-E acquisition rate was similar in individuals not exposed to nitrofurantoin in nitrofurantoin exposed households to that in households not exposed to nitrofurantoin (HR=0.85, 95%CrI=[0.34,1.85]), Figure 2). Higher FQR-E acquisition rates were associated with households from Lodz (HR=2.54, 95% CrI=[1.05, 6.03]) relative to households from Antwerp, but there were no clear differences in rates of FQR-E acquisition in households from Geneva compared to Antwerp (HR=0.57, 95%CrI=[0.22,1.34], Figure 2B(i)).

### Duration of FQR-E carriage within households

Mean baseline rate of FQR-E clearance at the household level was 0.07 (95%CrI=[0.04,0.18]) cases per day. Exposure to nitrofurantoin (HR=0.43, 95%CrI=[0.19,0.95]) or fluoroquinolone (HR=0.71, 95%[0.20, 1.44]), were both negatively associated with the household clearance rate (Figure 2), and thus prolonged duration of carriage. We assumed that FQR-E clearance in a household member was independent of the risk factors of other members within the household, hence we did not anticipate any indirect effects of nitrofurantoin or fluoroquinolone exposure on within household FQR-E clearance. We estimated the total household duration of carriage as the sum of all times spent in any state where at least one individual was colonised. For the two-member households, we estimated a baseline median total duration of carriage of 31 days. The median total duration of carriage in three-member households was 55 days.

## Discussion

Findings of this study indicate that fluoroquinolone and nitrofurantoin exposure are associated with higher and lower rates of FQR-E acquisition, respectively, while exposure to either class of antimicrobial agents is associated with prolonged duration of FQR-E carriage, both at individual and household levels. The study further shows that community rates of FQR-E transmission are higher within households than between households. The association of fluoroquinolone with FQR-E acquisition was more pronounced at the household level than at the individual level.

A particular strength of this study is in our use of multi-state Markov models to fit to the panel data. This allowed us to fit mechanistic transmission models accounting for interval censored and auto-correlated data while including exposures of interest (most notably, antimicrobial treatment), giving us the ability to characterise the partially observed state evolution of households and individuals over time and to quantify the association of these dynamics with antimicrobial exposure. An important limitation of the study is that we had to treat FQR-E as a homogeneous group of bacteria. It is likely that some of the transmission events involved different species of Enterobacteriaceae or genetically unrelated strains of the same species, however the data lacked the resolution needed to make these distinctions. High resolution characterisation of bacteria such as through the use of whole genome sequence data would allow for a higher resolution analysis of these transmission dynamics.

Acquisition and carriage dynamics of drug-resistant bacteria remain poorly described especially within households, where person-to-person transmissions are likely to occur due to frequent direct and indirect contacts between members. Whilst the individual-level positive association between acquisition of FQR-E and fluoroquinolone exposure are in agreement with previous findings [15, 24], the negative association we found between FQR-E acquisition and exposure to nitrofurantoin was not previously reported [15]. With antimicrobial consumption identified as a major driving factor for the emergence and spread of AMR, there is an urgent need for treatment strategies that optimise use of available antimicrobial agents to prevent or limit development of AMR [15, 25]. However, development and implementation of such strategies is challenging due to lack of evidence to guide agent selection in a way that maintains treatment effectiveness without selecting for increased resistance [15]. FQR has been shown to be frequently co-expressed with resistance to a number of other antimicrobial agents such as beta-lactams, sulphamexazole-trimenthoprim and aminoglycosides [26, 11]. Resistance to nitrofurantoin, on the other hand, tends to emerge slowly, irrespective of its usage, and it is rarely associated with resistance to other antimicrobials [11, 27]. This makes its association with reduced FQR-E acquisition rates interesting. A previous study reported a change in trend in FQR *E. coli*, from a steady increase during a period when fluoroquinolone was the recommended first-line agent for acute uncomplicated cystitis, to a declining trend after nitrofurantoin became the recommended first-line agent[25]. If the relationship between FQR-E acquisition and fluoroquinolone or nitrofurantoin is causal, then the findings of our study provide further evidence to support nitrofurantoin use in place of fluoroquinolone where possible, as an effective intervention that would reduce FQR-E spread. This would help to preserve fluoroquinolones for more severe infections and those infections with limited available antimicrobial therapeutic options.

Carriage is a prerequisite for infection and prolonged carriage duration of drug-resistant bacteria is likely to lead to a higher risk of drug-resistant infection in the carrier as well as causing more onward transmission of the bacteria to other persons [28]. In this study, we have shown that exposure to fluoroquinolone, nitrofurantoin and other antimicrobial agents is associated with prolonged duration of carriage. Our estimated individual level duration of carriage is consistent with previous estimates of FQR-E carriage duration of one month or less, and an association between prolonged duration of FQR-E carriage and fluoroquinolone exposure has also been previously reported [29, 30]. The positive association of antimicrobial exposure with duration of carriage might be attributed to the effect of antimicrobials on the gut microbiota. Antimicrobial exposure may alter the gut environment and reduce gut microbiota diversity leading to within host proliferation of drug-resistant bacteria and reduction of the gut’s ability to clear colonisation [31]. Available evidence indicates that fluoroquinolone and nitrofurantoin co-resistance is uncommon [27]. This suggests that FQR-E persistence despite nitrofurantoin exposure is unlikely to be due to acquired nitrofurantoin resistance traits, but could be explained by either FQR-E being able to survive nitrofurantoin exposure in the gut through other mechanisms such as antimicrobial tolerance or persistence, or that with less diversity, rate of turnover of colonising bacteria is lower.

Our investigation of household transmission dynamics revealed higher rates of FQR-E acquisition sourced from within than between households, indicating that person-to-person contacts within households (either directly or indirectly) are an important route of FQR-E transmission. Often, strategies for controlling spread of multi-drug resistant bacteria such as hand hygiene, antimicrobial stewardship or isolation of drug-resistant bacteria carriers are enforced and implemented within hospitals. The findings of this study, as with previous studies reporting high within household transmission rates [32], highlight the need for interventions that aim to limit within household transmission of FQR-E, especially when some household member(s) are potential carriers of FQR-E, for example, those that have just been discharged from hospitals.

It is also interesting to note that individuals and households from Lodz, despite having higher rates of FQR-E acquisition, had shorter duration of carriage than individuals from Antwerp and Geneva. The explanation to this is not immediately clear, but might reflect differences in species or strain composition of FQR-E at the different sites. It has previously been shown that duration of carriage can vary by species and strain of drug-resistant bacteria [29, 30]. With species or clonal classification of our isolates not available, we were unable to determine how bacteria species or specific clones of our isolates influenced the duration of carriage in this study.

This study has identified within household contacts as an important route for FQR-E transmission and highlights the need for considering household interventions to control FQR-E spread in communities. While nitrofurantoin use instead of fluoroquinolone may offer some protective effect against FQR-E colonisation, this benefit may have as a cost, prolonged FQR-E carriage when individuals exposed to the nitrofurantoin are already colonised by FQR-E.

## Methods

### Study design and data

The SATURN multinational cohort study has been described in detail elsewhere [15]. Briefly, ambulatory patients were recruited from three European sites including the General Practice Hospital Networks of Antwerp (Belgium) and Lodz (Poland), and the ambulatory clinics at the University of Geneva Hospitals (Switzerland). Study participants comprised of an exposed group, a convenience sample of patients with UTI who required antimicrobial treatment; and a non-exposed group consisting of patients presenting to the same clinics but not requiring antimicrobial treatment. To be eligible for recruitment, participants had to be at least 18 years old and living with at least one other person in the same household. For each index patient, one to three household members were recruited. Patients were not included in the study if they were being treated with systemic antimicrobial agents, had been hospitalised within 30 days prior to enrolment, were residents in a long-term care facility, were on a urinary catheter at the time of enrolment, or had renal transplant or renal replacement therapy. All participants provided written informed consent. Each participant completed a self-administered questionnaire at baseline and with each sample collection to record exposures to antimicrobial agents. Study participants provided three faecal samples: i) at baseline; ii) at completion of treatment with antimicrobial agents for the exposed group or seven to ten days after first sample for those not on treatment with antimicrobial agents; and iii) at 28 days after the second sample (Figure 3). Microbiological analyses were carried out on the faecal samples to screen for presence of FQR organisms and species confirmation.

**Figure 3:**
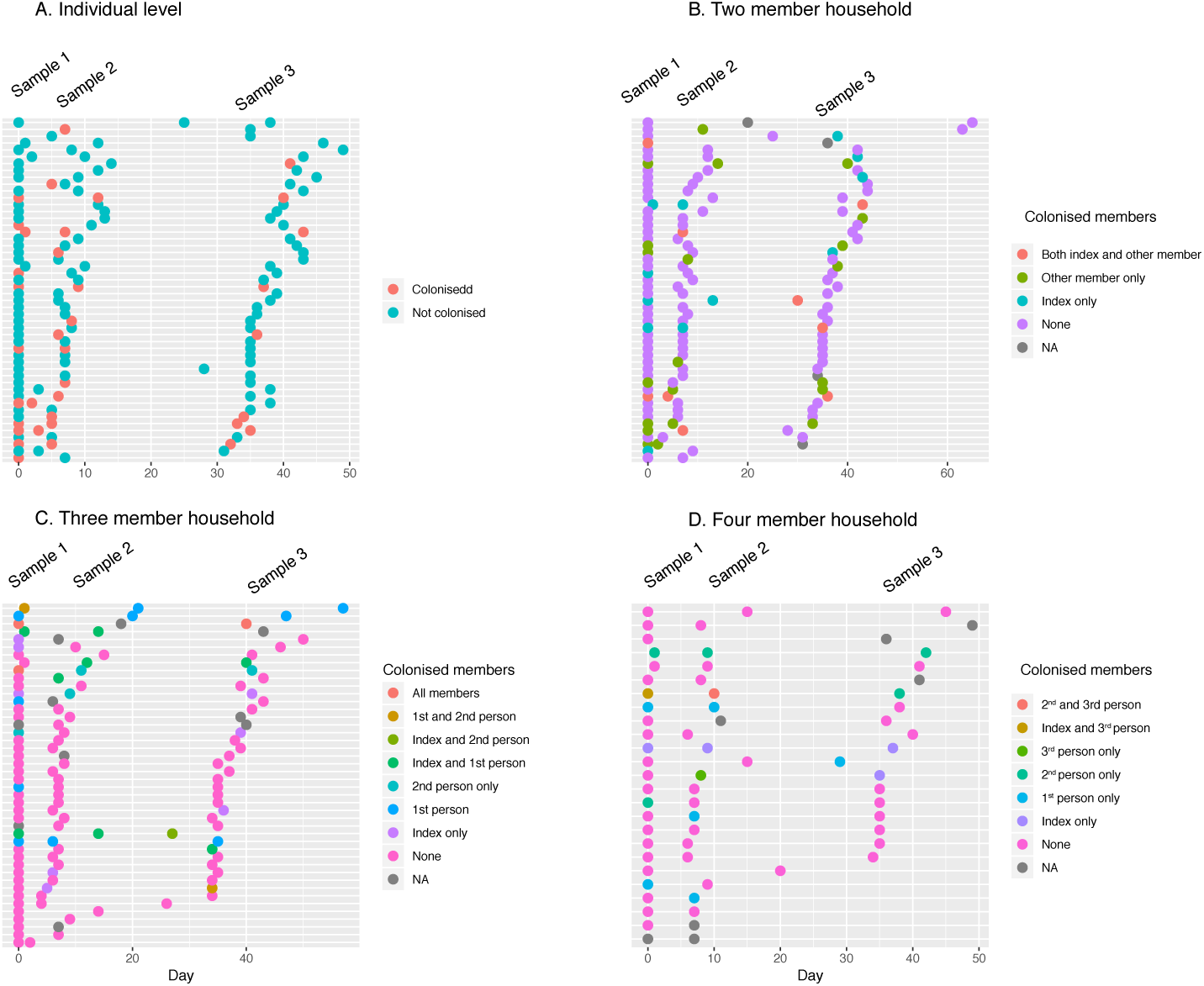
Fluoroquinolone resistant *E. coli* (FQR-E) colonisation outcomes of stool samples by individuals and households. Each row of dots in panel A depicts stool samples from an individual at the corresponding time and their FQR-E status (poistive or negative). In panels B-D, a row depicts the colonisation status of two-member, three-member or four-member households, respectively, defined by the number of colonised individuals within the household. Panels A and B only show subsets of individuals and households, for easy visualisation.

### Modelling transmission dynamics

We modelled acquisition and carriage dynamics of FQR-E at individual and household levels using continuous time Markov Models, implemented within the Bayesian framework. At the individual level, we first used dichotomous phenotypic data for fluoroquinolone susceptibility (sensitive or non-sensitive) and considered all participants, including index patients and their corresponding household members, to belong to one of two states: not colonised with FQR-E (state 1) or colonised with FQR-E (state 2) at the sampled time points (Figure 4a). Secondly, we used semi-quantitative data based on the density of FQR-E detected in the samples. Here, at any time point, individuals were considered to belong to one of three states: no FQR-E colonisation (state 1), low-density FQR-E colonisation (state 2), or high density colonisation (state 3) (Figure 4b). We then developed separate but related models for households with different sizes. An *n−*member sampled household had a total of *K* = 2^*n*^ possible states and households transitioned from one state to another depending on the colonisation status of each household member (Figure 4C).

**Figure 4:**
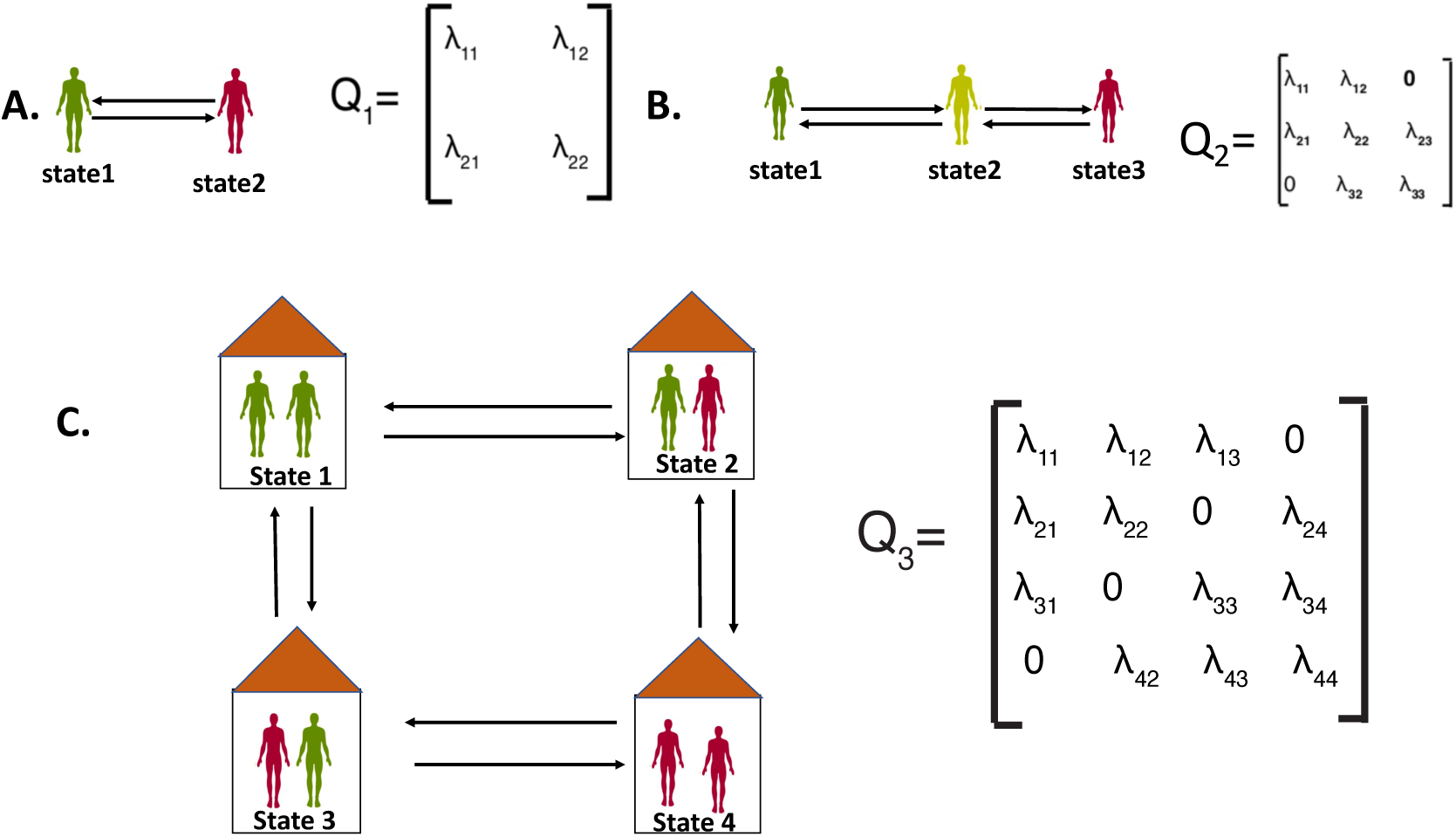
Conceptual multi-state Markov models for acquisition and carriage dynamics of fluoroquinolone resistant Enterobacteriaceae (FQR-E). (A) A two-state individual level model with a 2 × 2 transition matrix *Q*_1_. Individuals transition from “state 1” (non colonised) to “state 2” (colonised) at a rate *λ*_12_ per day and from “state 2” to “state 1” (cleared of FQR-E colonisation), at a rate *λ*_21_. Individuals remain in state 1 or state 2 at daily rates of *λ*_11_ and *λ*_22_, respectively. (B) Three-states (“no colonisation”, “low-density colonisation” and “high density colonisation”) individual-level model with a 3 × 3 transition matrix *Q*_2_. Individuals transition from state 1 to state 2 and from state 2 to state 3 at daily rates of *λ*_12_ and *λ*_23_, respectively; and clear from high density colonisation (state 3) to low density colonisation (state 2) at the rate *λ*_32_. Individuals clear from any detectable colonisation (from state 2 to state 1) at the rate *λ*_21_. Direct transitions between states 1 and 3 are not allowed. Results of the three-state household model are presented in Supplementary. (C) Four-state, two-member household model with a 4 × 4 transition matrix *Q*_3_. For this model, a household is in state 1 if neither of the members are colonised; in state 2 if only the index patient is colonised; in state 3 if the non-index member is colonised and the index patient is not; and in state 4 if both the index patient and the non-index household member are colonised. Households transition from state 1 to state 2, state 1 to state 3, state 2 to state 4 and state 3 to state 4 (FQR-E acquisitions) at rates *λ*_12_, *λ*_13_, *λ*_24_,and *λ*_34_ per day, respectively; and they clear from state 2 to state 1, state 3 to state 1, state 4 to state 2 and state 4 to state 3 at rates *λ*_21_, *λ*_31_, *λ*_42_, and *λ*_43_. No more than one instantaneous transitions are allowed hence, no direct transitions between states 1 and 4 and states 2 and 3.

In all individual-level models, we assumed that transition of participants between states was a first order Markov process governed, for individual *i*, by an intensity matrix *Q*_*i*_ where element *q*_*rs,i*_ of *Q*_*i*_ is the rate at which subject *i* transitions from state *r* to state *s*. Given a set of covariates {*x*_*i*,1_, *x*_*i*,2_, …, *x*_*i,D*_}, we modelled the daily rate of an individual *i* transitioning from state *r* to state *s* (*r* ≠ *s*) as

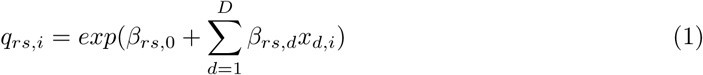

We included exposure to antimicrobial agents (nitrofurantoin and fluoroquinolone), location, travel to AMR endemic regions and use of any antimicrobial in the previous year as covariates to the two and three states individual models.

Colonisations within a household could either be from the wider community or from a within household source. Given *λ*_*i,h,C*_, the rate of an individual *i* in household *h* being colonised from contact in the wider community (between household transmission) and *λ*_*i,h,H*_ the rate of individual *i* in household *h* being colonised from a contact within the household (within household transmission), we modelled the overall household FQR-E acquisition rate for an individual *i* in household *h* as

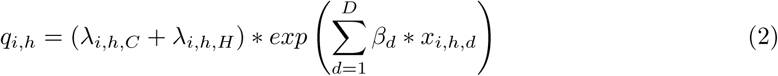

where {*x*_*i,h*,1_, *x*_*i,h*,2_, …, *x*_*i,h,D*_} are the covariates for individual *i* in household *h*. *λ*_*i,h,H*_ = 0, if the transition was from a state where no household member was already colonised.

We included exposure to antimicrobial agents (nitrofurantoin or fluoroquinolone), and study location as covariates. The household was considered to have had antimicrobial exposure if any member was recorded as having used antimicrobial agents during the follow up period. To account for this, we included in the household acquisition model parameters *β*_1,*I*_ and *β*_*2,I*_ (the direct effects of nitrofurantoin and fluoroquinolone exposure) if the transition from state *r* to state *s* involved colonisation of the index household member. We included parameters *β*_1,*NI*_ and *β*_*2,NI*_ (the indirect effects of nitrofurantoin and fluoroquinolone exposure) if the the transition from state *r* to state *s* involved colonisation of a non-index household member.

The diagonal entries of the transition matrix, *q*_*rr*_, were given as:

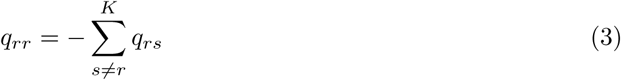

We then estimated the overall duration of carriage for a household of size *n, D*_*n*_ as the sum of times spent in all states of at least one colonised household member, i.e.

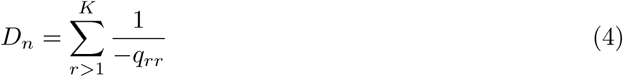

The data generating process of observation *y*_*it*+*δt*_ *∈* {0, 1, …, *K*} for individual *i* at sampling time *t* + *δt* followed a categorical distribution and hence the likelihood function:

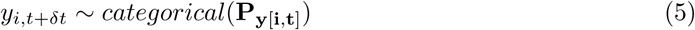

*i* at sampling time *t* + *δt*

Where **P** is a probability transition matrix and *δt* is the time-interval between two consecutive sampling time-points such that

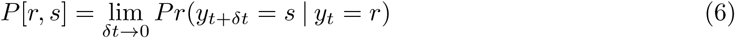

We assumed that the transition probabilities in **P** are differentiable at any time *t* and that the Markov chain governed by these transition probabilities is time-homogeneous, hence satisfying the Kolmogrov forward equation:

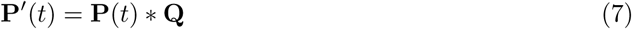

which has the matrix exponential solution **P**(*t*) = *exp*(*t***Q**) [33] and we used this relationship to convert the intensity matrix to interval-censored daily probability transition matrices for the likelihood function.

We used weakly informative Gaussian prior, normal(0,1) for all *β*s and uniform prior, uniform(0,10) for the between household and within household baseline rates, *λ*_*i,h,C*_ and *λ*_*i,h,H*_, respectively. We fit models to data using the Hamiltonian Markov Chain Monte Carlo implemented in Stan (version 2.17.3) within the R environment (v. 3.4.4). We assessed model convergence and performance by manual visualisation of the traceplots, which showed satisfactory convergence (Figure S1 and Figure S2) and the Gelman-Rubin convergence statistic (R.hat). We compared performance of the two-states and three-states individual level models by assessing how well parameter estimates from the two models simulated the observed data. We fitted the household model to two and three member household data only. There were very few four-member households and extension of the model to these households resulted in no numerical solutions being obtained.

## Data Availability

All data referred to in the manuscript have been summarised and reported within the manuscript. Raw data and code used to analyse the data will be made available online.

## Acknowledgements

We thank members of SATURN WP3 study group (Caroline Brossier, Cécile Delémont, Begonã Martinez de Tejada, Gesuele Renzi, Jacques Schrenzel, Sylvie Van Bylen, Jonathan Vanbergen, Philip Koeck, Peter Leysen, Kim Vandercam, Jacques Kluijtmans, Agnieszka Borkiewicz, Frank Heyvaert, Nele Michels, Gerd Deswaef, Tine Beckx, Hadewijch Declerck, Katy Embrechts, Nina Verheyen, Tine Bauwens, Josefien Béghin, Liesbeth Verpooten, Katrien Bombeke, Tine Vandenabeele, Magdalena Muras, Joanna Świstak, Anna Wesolowska, Ewa Sterniczuk, Lidia Brzozowska, Krystyna Cichowska, Jolanta Szewczyk, Grażyna Krupińska, Honorata Błaszczyk, Urszula Stawińska, Maria Szyler, Marek Kasielski, Rafał Rydz, Agata Myszkowska, Lilianna Zėbrowska and Benedikt Huttner) and the SATURN study participants.

The original SATURN cohort study was supported by the European Commission under the 7th Framework Programme (FP7-HEALTH-2009-SINGLE STAGE-SATURN, contract 241796); and by the Methusalem grant of the Flemish government to Herman Goossens. PM was supported by the UK Medical Research Council (MRC-UK) JPI-AMR MODERN Project grant (MR/R004536/1) to BSC. AJS is supported by an Australian National Health and Medical Research Council Early Career Fellowship (GNT1141398). BSC was supported by the MRC-UK Senior Fellowship Grant (MR/K006924/1).

