## Supplementary material for "Acquisition and carriage dynamics of fluoroquinolone resistant Enterobacteriaceae at individual and household levels"

### Supplementary Results

#### Acquisition and carriage dynamics of FQR-E using a three states individual level model

We also considered the density of FQR-E in stools with an individual-level model with three states: not colonised, low density colonisation, and high density colonisation. The baseline rate of transitioning from no colonisation to low density colonisation was 0.4 (95% CrI=[0.10,1.54]); the colonisation density increased to high from low at a baseline rate of 0.07 (95% CrI=[0.04,0.14]) per day. Individuals transitioned from the high density colonisation state to the low density colonisation state at a baseline rate of 0.06 (95%CrI=[0.03,0.10]) per day and were cleared of any detectable colonisation at a daily rate of 3.37 , although this was associated with a high amount of uncertainty (95% CrI=[ 0.83, 13.04]).

### Supplementary Figures

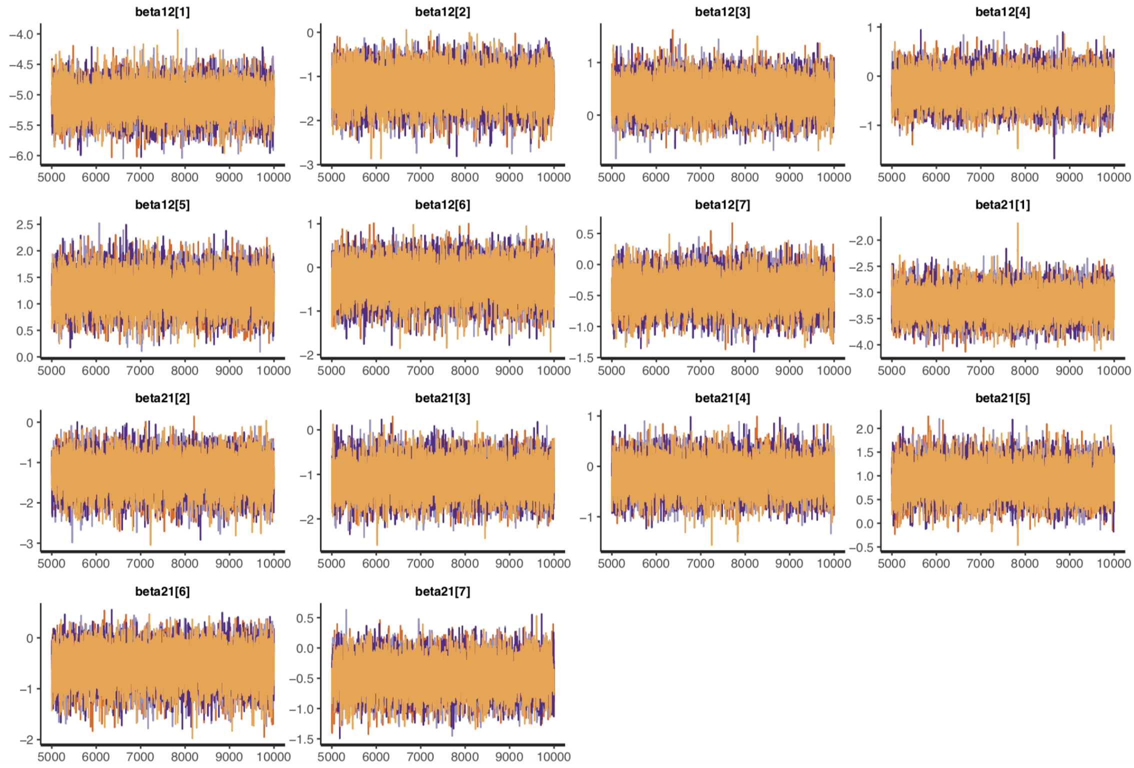


Figure S1: Traceplots for the Hamiltonian Markov chains, showing sampling behaviour and mixing across chains under the two-states individual level model

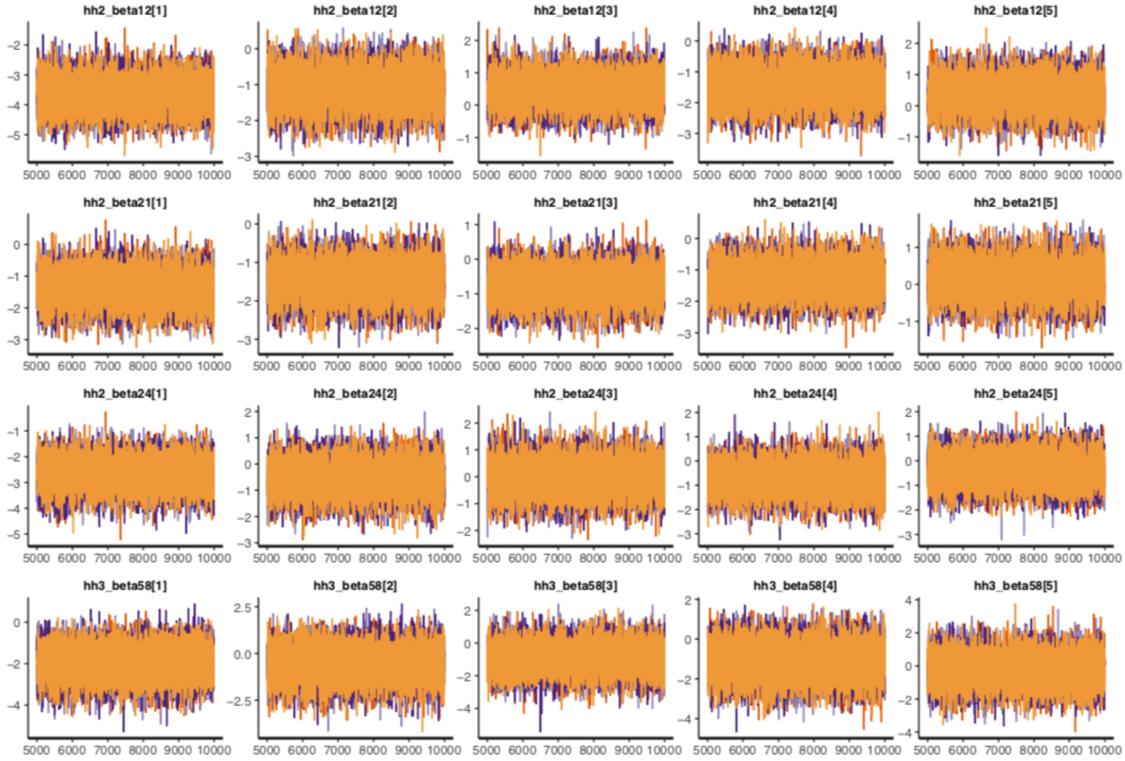


Figure S2: Traceplots for the Hamiltonian Markov chains, showing parameter value sampling behaviour and mixing across chains under the household level transmission model.
